# Participant experience of Scottish Ballet’s dance-based long COVID support programme: A mixed methods study

**DOI:** 10.1101/2024.10.28.24316302

**Authors:** Keir EJ Philip, Alexandra Burton, Adam Lewis, Sara C Buttery, Parris J Williams, Michael I Polkey, Daisy Fancourt, Nicholas S Hopkinson

## Abstract

**Objectives:** To explore participant experience and perceived impacts of an online dance-based long COVID support programme.

**Design:** Mixed-methods study using thematic analysis of semi-structured interviews and structured observations, and ordinal scale questionnaire responses.

**Setting:** Online, community-based, with participants in Scotland and England.

**Participants:** 26 people with self-reported long COVID who participated in the final block of 10 sessions were invited to participate in this study. 13 of these consented, 12 female, 7 White British, 10 English as first language, mean(range) age 57yrs (38-77), symptom duration 27months (17 – 35), and live sessions attended 9.2 (4-17). Two dance leaders also participated in the study.

**Interventions:** An online dance-based long COVID support programme of live sessions, provided in morning and afternoon slots, 30 to 45 minute duration, and online materials to promote wellbeing and self-management strategies, developed by Scottish Ballet, people with long COVID and healthcare professionals. The block of 10 sessions, from which participants were recruited, ran in September-November 2023. Potential participants were required to confirm they were well enough to participate in the programme.

**Results:** Responses to closed questions regarding self-reported impacts demonstrated perceived physical and mental health improvements, the programme surpassed participant expectations, and participants would recommend the programme. Qualitatively, we identified four themes 1. Improved experience of symptoms; 2. Increased confidence regarding movement and physical activity; 3. Feeling calm and refreshed; 4: Valuing time spent in a safe space. Facilitators of impact related to content, logistics, and delivery. Few barriers were described. Dance leaders’ responses aligned with those of programme participants.

**Conclusions:** A remotely delivered dance-based intervention for supporting people with long COVID is feasible, with participants consistently reporting that they found it enjoyable and beneficial to their health and wellbeing. Further research to assess impact on clinically validated measures is required.

**Strengths and Limitations of the study:**

- This is the first known study to explore participant experience in a dance-based support programme for people with long COVID.
- The study provides insights into participant perspectives on their experience of movement and ways it might be incorporated into long COVID management.
- Small number of participants limits generalisability of the conclusions.
- Participants were self-selecting both in accessing the program and the evaluation described here.
- Further research is required to assess impact on clinically validated outcome measures.

## Introduction

Post-acute COVID-19 syndrome (‘long COVID’), is an often severe and disabling multisystem condition, characterised by persistent symptoms following infection with the virus Severe Acute Respiratory Syndrome Coronavirus 2 (SARS-CoV-2) [1–3]. Around 65 million people have been affected worldwide [4], and an estimated 1.9 million people (2.9% of the population) had long COVID in the UK in March 2023 [5]. As such, long COVID is having extensive health, societal and economic impacts [6–8]. Work is ongoing to understand the pathophysiological processes involved. These appear to include direct tissue damage from the virus, immune system abnormalities, chronic inflammation, and post-critical illness sequalae [4, 9]. Long COVID causes biological, psychological, and social impacts, which interact with one another leading to reduced quality of life [3, 10, 11]. A very wide range of symptoms have been reported due to the various organ systems that can be involved, which can vary between individuals and over time [12, 13]. Common features include extreme fatigue, cognitive processing difficulties, labile heart rate and blood pressure, and breathlessness [8, 9, 14, 15].

There are currently limited evidence-based interventions for long COVID management. However, clinical guidelines suggest personalised and holistic approaches to support recovery [16, 17]. Participatory arts engagement can improve physical and mental health in the general population and in individuals with specific health conditions [18–22]. Previous theoretical work has conceptualised the component parts of arts interventions as ‘active ingredients’ consisting of the design features of the activity as well as the social and physical environment in which the activity is delivered [23]. These can then activate diverse mechanisms of action – psychological, biological, social and behavioural processes leading to health-related outcomes[24]. Singing and breathwork approaches have been shown to improve breathlessness in other conditions, through changing the way in which people breath (posture and breathing pattern), and, through improving the experience of breathlessness through lowering associated anxiety levels [25–27]. Dance-based approaches can improve anxiety and depression through positive experiences of social connection, body experience and perception, and spending time participating in an enjoyable activity [28–32]. Dance-based approaches have also been shown to improve physical function and capacity, through exercise training impacts [28, 33–36]. Additionally, the collective and social aspects of certain participatory arts interventions have been used effectively to reduce feelings of social isolation and loneliness [30, 31, 37]. The limited research that exists on online dance-based interventions suggests potential for similar impacts [38, 39].

Current examples of arts-based interventions being developed for long COVID include the English National Opera’s (ENO) Breathe programme, which is an online breathing and wellbeing programme that uses singing techniques. A randomised controlled trial found that it improved aspects of health related quality of life and breathlessness for people long COVID and breathlessness [40]. Another example is a theatre-based wellness programme for people with long COVID developed by The Old Vic. This was found to be acceptable and feasible, with exploratory outcome measures suggesting possible improvements in general health and chronic fatigue symptoms[40]. Such approaches do not reverse physical damage caused by the virus. Rather, they attempt to support psychosocial aspects of living with the condition to help people manage physical symptoms and to change the way that they experience them.

Scottish Ballet worked with people with long COVID, and healthcare professionals to develop an online dance-based long COVID support programme of weekly live sessions and online materials to promote wellbeing and self-management strategies. Programme delivery took place during 2023. The aim of this study was to explore participant experience and perceived impacts, barriers and facilitators, of this programme.

## Methods

### Research design

#### Intervention

Scottish Ballet’s long COVID support programme is the newest of Scottish Ballet’s Dance Health programmes. The pilot programme was delivered between March and October 2023 and was funded by The Rayne Foundation.

#### Programme Content

The programme consisted of weekly online sessions, online resources, and regular contact and support for the participants from the Dance Health programme team. The content was delivered by highly experienced dance for health practitioners from Scottish Ballet.

##### Weekly sessions

Sessions took place live, online, lasting 30-45 minutes, using the video conferencing platform Zoom. A block of 18 weeks was run in the spring/summer 2023, and ten weeks in autumn 2023. A morning and afternoon session were offered during blocks and people could pick which sessions they wanted to attend from any of those offered.

##### Online resources

Online resources were provided to complement the weekly live sessions. Each online resource could be used as a standalone session or in conjunction with others depending on the desires of the individual participant. Each resource was composed of:

- Directions to help the participant ‘check in’ with their body and their emotional state
- Gentle movement for strength and mobility
- Breathing exercises
- Guidance to help participants manage their energy and fatigue
- Guided dance-based movements with seated, standing, and lying down options – providing options for the participant to choose what felt good for them
- Clear safety guidelines
- Specially composed music

##### Contact between participants and the Dance Health team

Participants received regular emails and contact from the session leaders. Emails encouraged participation and resent links to resources and live online sessions to facilitate access.

##### Programme development

The pilot programme content was created in collaborative partnership with people living with long COVID, long COVID researchers, and healthcare professionals with expertise in long COVID and experience providing care for this group. The content was developed based on information and feedback during their initial research and intervention development phase. This process played an important role in creating a programme that was reflective of participants’ needs, abilities, and preferences. Specific adaptations related to fatigue, brain-fog, and language were vital. For example, the videos were kept reasonably short and low intensity, in terms of speed and size of movements, due to potential issues related to fatigue and exertion related symptom exacerbation which are common symptoms. An example of the videos can be accessed here: https://youtu.be/CEV4GptTBP8. The programme was developed with the aim of providing movement resources and tools to help people with long COVID to self-manage symptoms and build resilience. The programme focused on improving posture, alignment and mobility in addition to using creative tasks to build confidence and self-expression. We have included a completed INgredients iN ArTs in hEalth (INNATE) Framework checklist[23] in the supplementary material which documents the key project, people and environment details of the programme.

This mixed methods study took place as a component of a larger evaluation of the pilot programme. The wider evaluation encompasses an internal evaluation conducted by Scottish Ballet which includes more information on the development of the programme; logistical considerations of running the programme; and potential future developments.

##### Evolution during the pilot phase

The original planned programme was to conduct the weekly live sessions in person in a dance studio, which would have been followed by a short Social Café at Scottish Ballet, with additional creative digital resources. However, following consultation with people with lived experience of long COVID and healthcare professionals, it became clear that in person sessions would not be feasible for many people, as the very act of getting to the sessions and back again, would have been excessively demanding, before even considering the energy and exertion required for the sessions. Therefore, the decision to deliver the programme exclusively online was based on participant preference and adaptations related to long COVID related limitations. These changes also necessitated adaptation of this evaluation and the related ethics approvals. The initial plan was to complete a mixed methods assessment using pre and post intervention assessment of validated questionnaires as well as the qualitative data analysis presented here (see protocol attached). It was also apparent that the number of study participants was likely to be insufficient to sufficiently power quantitative analyses and that a large battery of measures may worsen participant fatigue, which is an important feasibility consideration. Therefore, to reduce overburdening with assessment, we decided to focus on gathering people’s experiences of the programme qualitatively and via a short questionnaire at the end of the interview. A substantial amendment to the ethical approval was granted.

##### Programme Recruitment

Scottish Ballet advertised the pilot through healthcare professionals involved in the development of the programme and incubation phase, existing long COVID groups and word of mouth. Information and contact details were provided on the Scottish Ballet’s website. Before beginning the programme, potential participants were required to confirm that, as far as they were aware, participating in the programme would not pose any significant risk to their health. For example, if they experienced episodes of chest pain or collapse on minimal exertion, they would not be eligible to participate.

##### Research Procedures

###### Study recruitment

69 people participated in the programme. 26 people participated in the final block of 10 sessions and were invited to participate in this study. People who had not participated in the final block were not invited as it was felt the time since participation might impact their responses. Programme participants were approached in the 2-4 weeks before the programme started to see if they would like to participate in the evaluation. The artists leading the sessions were approached to take part in an interview at the end of the programme. All potential study participants were provided with information on the study by Scottish Ballet staff and provided with contact information for the study team if they would like more information. Individuals who contacted the research team and were participating in the programme, were then invited to participate in the study. The study aims and objectives were explained and Participant Information Sheet (PIS) provided, with at least 48 hours to read, consider and ask questions. Written informed consent was obtained from all participants. Participation in the long COVID support programme did not require participation in this study. There were no exclusion criteria for participation in this study.

###### Data collection

Data were collected using semi-structured interviews with participants at the end of the programme. These took place via a video conferencing application and were conducted by KEJP (PhD) who is a male Clinical Lecturer and medical doctor, trained and experienced in conducting qualitative interviews with people with long term health conditions. A relationship was established prior to the interviews by introducing himself to the group in one of their sessions, describing the research, reasons for conducting it, and answering questions. KEJP has a PhD investigating arts-in-health interventions. A topic guide was used (see supplementary material) which was developed by reviewing related research including arts-in-health interventions for other conditions and long COVID. Basic demographic data was also collected including age, gender, ethnicity, and self-reported medical conditions. Participants were also asked to describe the main symptoms they were experiencing. Audio recordings were transcribed verbatim. One participant requested that the interview was not recorded but was happy for note taking, so this approach was employed.

Closed questions were also included at the end of the interview, focusing on key questions of interest related to perceived change of components of health, attribution to participation in the programme, achievement of expectations, and whether they would recommend the programme to other people with long COVID. The responses to these questions are provided in Figure 1 and Figure 2.

**Figure 1:**
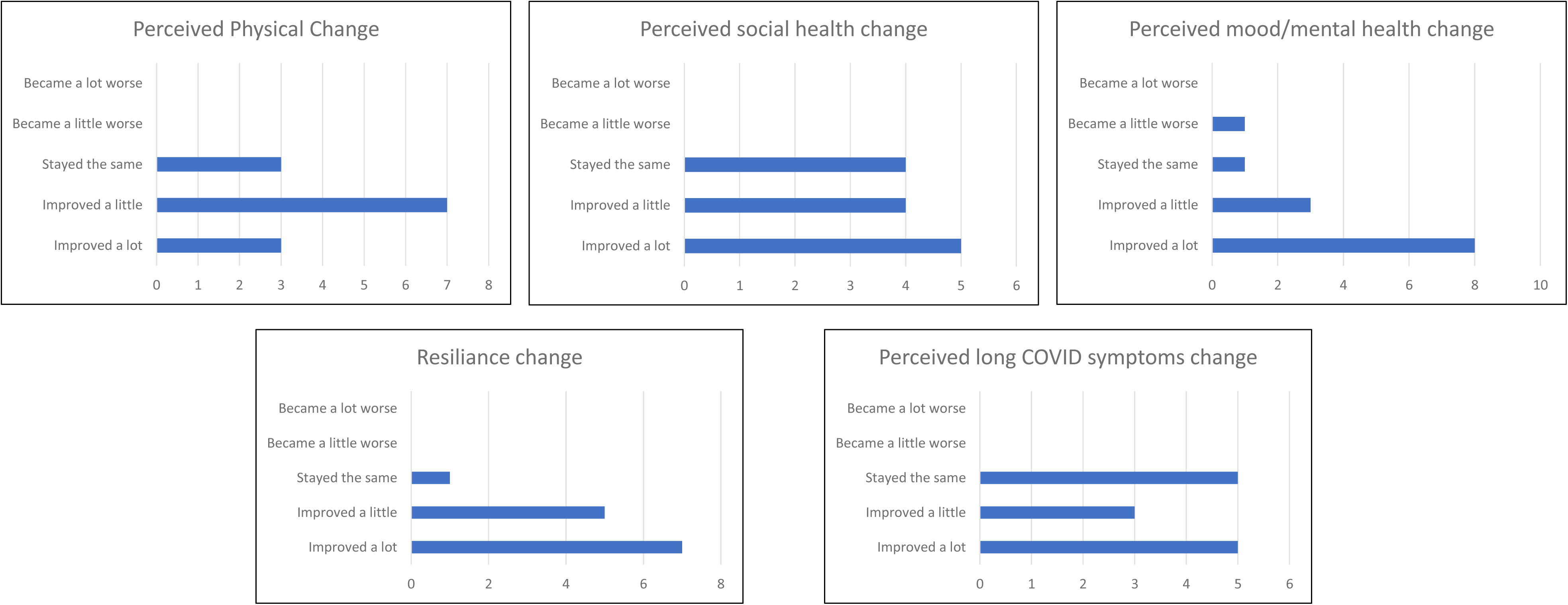
Multi-panel of graphs of perceived health impacts related to participation

**Figure 2:**
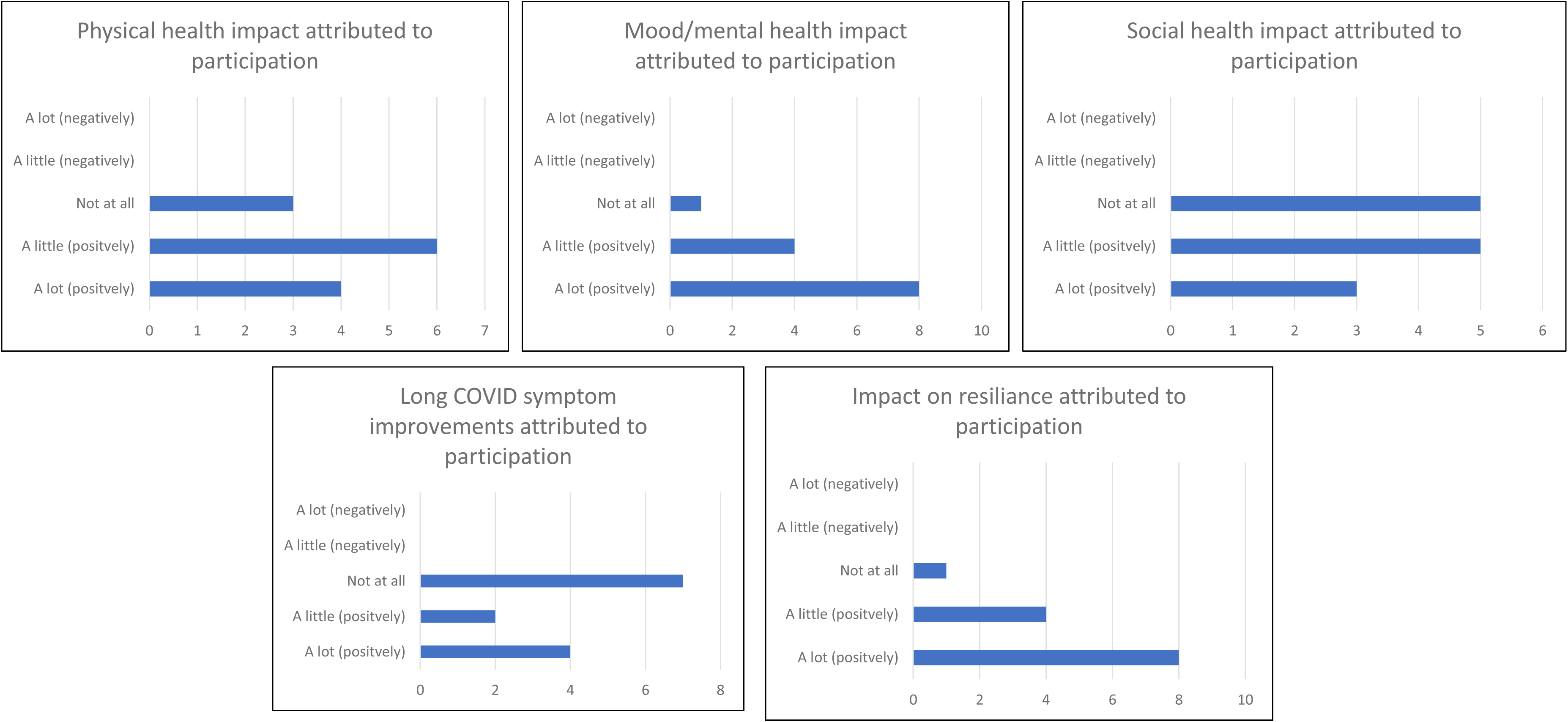
Multi-panel of graphs of amount of change in health attributed to participation

KEJP and AL also conducted structured observations of live sessions and online resources to provide contextualisation for analysing the interviews. These were made by observing sessions, but not participating, under predefined headings, of session content, participant interactions, participant movement, psychological impressions, and other. This allowed for a better understanding of the structure and content of the programme and interpretation of interview and other data.

All data collection was completed within four weeks after the programme finished. Adverse event monitoring relating to participating in the programme was monitored by Scottish Ballet. Although this was not part of the current study, potential adverse events related to participation was a topic of interest explored during the interviews as it relates to impacts and experience.

### Data analysis

We used reflexive thematic analysis[41] within a critical realist paradigm. Our approach to coding took a predominantly inductive and semantic approach, directed by the context while reflecting the explicit content of what participants said. The structured observations provided further context for the data analysis. Interview transcripts and observation notes were read and reread for familiarisation, before being uploaded into NVivo for coding. First cycle codes were created by initial open coding, which were then refined. All transcripts were assessed in this way by KEJP, with secondary coding of manuscripts split between AL, AB, SCB, PJW. The researchers then discussed their coding, grouping conceptually similar codes, and developed themes, with reference to the context and addressing the research questions and topics. Quantitative data from closed interview questions was analysed descriptively in Excel.

### Ethical approvals

Ethical approval for this study was granted by the Imperial College London Research Ethics Committee (ICREC Reference number: 6533422) on 20/03/23) which was conducted in accordance with the recommendations for physicians involved in research on human subjects adopted by the 18th World Medical Assembly, Helsinki 1964 and later revisions. Signed participant consent was obtained.

### Patient and Public Involvement

The focus of the current study was on the experience of participants with lived experience of long COVID, and people with long COVID were throughout the intervention development, helping to shape the content and duration of the programme. A draft of this manuscript has been reviewed by all study participants to sense check the content.

## Results

### Participants

Of the 26 people who participated in the final block of 10 sessions, who were invited to participate in this study, 13 contacted the research team to express interest in participation and all took part in the study. Reasons for not contacting the study team are unknown for the remaining 13 programme participants as there was no direct contact between the researchers and potential participants unless participants initiated contact. 12/13 (92%) were female, the majority self-identified as British/Scottish and had English as a first language. Five of the participants were currently working, eight were not. Of those not working, four were not working due to ill health, and four were retired (one of whom retired early due to ill health). Participants reported various types of occupation, past and present, including education, health, and creative industries. Those reporting comorbidities included ‘allergies’, asthma, depression and osteoarthritis. Mean duration of long COVID symptoms for the group was 27 months (range 17 to 35 months). Mean interview duration was 55.50 minutes. Data saturation was felt to have been met. More details on participant characteristics are provided in Table 1.

**Table 1:** Participant characteristics.

|  |  |
| --- | --- |
| Age (years) | mean 57 (range = 38 – 77) |
| Gender |  |
| Male | 1 |
| Female | 12 |
| Ethnicity |  |
| White British/Scottish | 7 |
| White European | 4 |
| European of other ethnicity | 2 |
| English as first language | 10 |
| Currently working | 5 |
| Not currently working | 8 |
| Living alone | 6 |
| Living with other medical conditions (comorbidities) | 4 |
| Duration of long COVID symptoms | 27 months (range 17 to 35 months) |
| Admitted to intensive care during acute COVID-19 | 1 |
| Admission to hospital during acute COVID-19 | 3 |
| Home monitoring during acute COVID-19 | 1 |

Participants attended a mean of 9.2 of the Scottish Ballet live sessions (range 4 to 17). A maximum attendance of 18 sessions was possible as some participants attended the earlier sessions then continued into the final group block. Online videos (YouTube) had a total of 413 views. It was not possible to separate out the viewing data of online resources for the current studies’ participants and views from other dance participants. In addition to participating in the Scottish Ballet long COVID support programme, participants reported various other supportive approaches including physiotherapy, occupational therapy, antihistamines and cardiac medications. In addition to the 13 participants, two dance artists (session leaders) were also interviewed.

Many of the participants found out about the programme through word of mouth and through social groups related to the English National Opera Breathe project. This route had the benefit of personal peer recommendation from a trusted person with whom a degree of understanding might be assumed. Others were told by physiotherapists or long COVID clinics or services.

#### Reported impact of long COVID

All participants described how long COVID had severely impacted them in a wide range of ways.

> *‘I just couldn’t do anything. Couldn’t participate in life. Really, my, my life totally changed.’ Participant 4*

Most participants, prior to long COVID, were in good or very good health. The most commonly reported long COVID symptoms included extreme fatigue, brain-fog, muscle and joint pain, palpitations, abnormal skin sensations, and breathlessness. These symptoms had persisted for many months resulting in major disruption to their lives. Socialising became difficult for some and impossible for others. Some participants lost their jobs or had to drastically adapt working schedules. All reported challenges to activities of daily living in one way or another. Many reported a lack of understanding or empathy from friends, family and healthcare professionals, and a lack of effective treatments or management strategies. Participants commonly reported being members of peer support groups, often online, which were generally felt to be useful though some expressed frustration regarding the occasionally peculiar therapeutic suggestions of peers, and the perceived focus on illness rather than recovery.

Although there was substantial overlap in symptoms between participants, there was also a large amount of heterogeneity regarding symptoms, severity and extent of improvement overtime.

The main focus of this study was to explore participant experience and perceived impacts, barriers and facilitators of the programme. Here we describe the themes identified and use anonymised participant quotes to illustrate them.

##### Perceived health and wellbeing impacts of the programme

Overall, experiences of participants were highly positive, *‘I loved it.’ (Participant 4)* . All stated they would, or already had, recommended the programme to other people with long COVID. No specific adverse events were reported. Thematic analysis of responses identified four key, inter-related, themes relating to how the programme supported health and wellbeing: 1. Improved experience of symptoms through reconnection with self; 2. Increased confidence regarding movement and physical activity; 3. Feeling calm and refreshed; 4: Valuing time spent in a safe space.

##### Theme 1 Improved experience of symptoms

Participants reported that the sessions provided them with an opportunity to engage with their body and the symptoms experienced. Taking the time to focus on sensory experiences enabled a more nuanced understanding and considered response to symptoms

> *‘…an increased sense of body awareness, I would say… I think I’ve learned a lot already in the last two years about reading body signs and understanding a bit more. What kind of activities make symptoms flare or how to mitigate some of that through breathing through slowing down. I I think it’s it’s that relationship between knowing your body a bit better.’ (Participant 3).*

They described how the sessions provided an opportunity to reconnect with aspects of their identity that long COVID had taken away.

> *‘Thank you, Scottish Ballet, for sorting this out because I love dancing and it’s my thing and that’s that’s all I wanna do in life is dance. (Participant 2)’*

Long COVID had resulted in a constant focus on what participants could no longer do, while the programme focused on what they could do. This was clearly noted in the structured observations. Rather than concentrating on limiting themselves to avoid discomfort, they were able to gently explore their capabilities, reconnecting with positive aspects of themselves in a creative way.

> *‘I think maybe anyone that suddenly has an illness or especially with long COVID where you know medically there’s not that much and that can be done and just being engaged in a creative way is just really uplifting and it really helps with kind of managing how you think about the process and so for me, as someone who liked them and the whole dancing part of it, it really helps and even umm like I even like just seeing the dancers in the studio while while I’m at home. Like it makes me feel like I’m I’m doing something like like a dance class’ (Participant 5)*
>
> *‘No mention of long COVID – person rather than disease focused’ Structured observation*

Participants described various coping strategies and techniques that they had learnt during the programme that they used outside of the sessions to help manage symptoms. These focused on using movement or breathing mindfulness techniques to reconnect with themselves in the present. These approaches did not stop the symptoms occurring but gave them tools that they found useful. In doing so, they reduced the psychological and social impacts of the symptoms. Most participants were clear that although these tools were useful, they were by no means a cure.

> *‘I use some of the some of the movement and the stretching in the mornings and the brushing [movement] and the breathing techniques before I get going in the morning.’ (Participant 1)*
>
> *‘I have to say to [friends and family] I’m not better. But what I’m ultimately doing is managing better. There’s a very slight subtle difference there. So, I still experience the issues that I was facing shortly after contracting the illness, but have learned coping mechanisms… I don’t feel that I’m necessarily better, but I manage better. So, and this is one of those tools* which helps me manage better. they start to become just another tool that’s in the box. So, at any moment, being able to put a little recharge in for me, especially in a work setting, is vital.’ (Participant 13)

Many participants had some prior dance experience and saw dance as part of their pre-long COVID self. As such, dancing in the programme enabled them to reconnect with that part of themselves. Previous experience ranged from enjoying dancing with their family, to using dance in their occupation.

##### Theme 2 Increased confidence regarding movement and physical activity

Building confidence in movement and participating in physical activity was central to the perceived impacts for participants. They described how the sessions had enabled them to extend their range of movement (degrees through which joints can comfortably move), gradually participate for longer, and perhaps most importantly, to gain back some confidence in their physical abilities.

> *‘with the Scottish Ballet, with the movement, the gentle movement, it’s just, it’s really good. Now I know how to breathe properly, I can get my movements, sort of working well together.’ (Participant 2)*
>
> *‘Just doing some simple stretching when some days perhaps you can’t do very much, it just shows you that you can do it.’ (Participant 6)*.

The most common reasons cited for participating in the programme were a desire to include movement in their recovery, valuing creativity, and/or wanting to feel they had ‘tried everything’. This was the case for those with previous formal dance experience and without. For those participants, dance and movement were important components of self that they wanted to reconnect with.

> *‘wanting something that allowed for movement and to physically do something, to feel like you were doing something was really key for me.’ (Participant 1)*
>
> *‘Well, I knew that I had to do some form of movement.’ (Participant 4)*
>
> *‘I wanted to try and find something that meant I got a bit more movement back as well. (Participant 13)*

The flexible nature of the activities provided in the sessions were important facilitators in supporting the development of confidence in movement. Participants greatly valued being able to participate sitting or lying down and feeling free to do as little or as much as they felt appropriate at that particular time - an understanding of there being no specific ‘right way’ removed any pressure to ‘perform’ or extend beyond their comfort zone. This self-selected personalisation, in which participants maintained control of intensity during the sessions was noted in the structured observations, supporting the interview responses. The way in which the sessions were delivered allowed participants to pace themselves, which enabled many to be able to safely engage in movement.

A further facilitator of increased confidence in movement was experiencing positive rather than negative impacts on energy,

> *‘somehow even after those kind of long and challenging days, I found them kind of weirdly reenergizing or, you know, in a way, its kind of calm and gentle and slow enough so that I didn’t feel I would overexert’ (Participant 3)*

##### Theme 3 Feeling calm and refreshed

Participants were explicit about the sessions having positive impacts on their mental health through helping them feel calm and to relax.

> *‘It does make me feel much calmer. It does change my mood, especially after he and I were doing all those exercises, you know, with the breathing and calming all your breathing down and everything is good. It just it makes me feel happy.’ (Participant 1)*

A range of impacts on mood and mental health were reported, with calming and refreshing being the most commonly reported direct impacts of the sessions,

> *‘More centred, more rested, and refreshed’ (Participant 5)*
>
> *‘Just calm, just calm. Reset. There’s nothing else to go, and you don’t need to think about anything. Just take a bit of of a moment for yourself and so I do it.’ (Participant 13)*
>
> *‘Participants appear relaxed and completely engrossed in the moment’ Structured Observation*

The creative component of sessions was commonly mentioned as being important. The powerlessness of long COVID was a major cause of anxiety and distress, so being active and creative was seen as empowering, novel and clearly an important ‘ingredient for impact’ for many.

> *‘…you’ve got all of that, lovely, you know, imagination part of it as well, which is wonderful. That obviously you know it’s really calming, and it’s really stressful having long COVID.’ (Participant 1)*

The use of imagery helped individuals achieve a deeper level of psychological engagement with the sessions than would have otherwise been possible with simple movement instruction, taking them away from the stress of their illness. Many appeared surprised at how powerful the experience was,

> *‘the way that the ladies actually describing things that - leaves falling onto the ground or crushing and sand between your toes and things like that, it was an easy and I don’t know how they do that. Yeah, it’s it’s amazing. And it takes them to a different place of thinking as well. (Participant 4)’*

Participants expressed high levels of trust in Scottish Ballet which enabled participants to relax into the activities. The session leaders were felt to be highly empathetic, caring and considered in the way they delivered the sessions, all contributing to the ‘safe space’ (Theme 4) that underpinned the calming impacts.

##### Theme 4 Valuing time spent in a safe space

Core to perceived health and wellbeing impacts was experiencing the programme as a safe space. The key ingredient being that the Scottish Ballet team were seen as professional and knowledgeable. Participants appreciated that the session leaders had both expertise in dance for health but had also conducted a substantial amount of preparatory work and research, to inform the long COVID specific aspects of the programme. It was clear to participants that all aspects of the programme, down to the detail, were carefully designed through appropriate means. This expertise resulted in participants feeling safe under their instruction,

> *‘I really like about it is that you know they’re, you know, you have two, two teachers and then the the the musician someone that does the technical thing. But you know, they’re all dancers and artists themselves, and I think that really makes it different from, like, doing yoga or no more like gym, like exercises like, you know, they’re artists.’ (Participant 5)*

The safe space was also created through the people taking part having a shared lived experience. This decreased feelings of isolation and increased confidence.

> *‘part of it is like with the, you know, program is seeing other people. Umm, but you know have the same thing, and you know it can be quite isolating sitting by yourself at home all day, so it’s nice to to kind of know that you are not alone.’ (Participant 5)*
>
> *‘So this is it for me, as it has been another way of getting back to talking to people quite freely and openly. And as I say, as I said earlier, you know, close friends and family. When I’ve talked about what’s I’m going through, haven’t quite got it, you know. People in those sessions, you feel like you can just be a little bit more yourself. Say what’s on your mind. Say how you’re feeling or say nothing at all, but again, just as a social element and when you’re looking at 12 or 15 faces on a screen, it’s really nice. You know there’s… it’s just, it’s nice to know that you’re not just there on your own sometimes.’ (Participant 13)*

This allowed participants to feel more confident being themselves, as they felt they would be understood, without having to explain themselves. Being in a safe space helped participants to practice ‘being myself again’. This also tied back in with Theme 1, with the ingredient of being a ‘safe space’ supporting reconnection with self.

The sessions also included optional time after the programme session for people to talk. Some participants found this useful, while others did not, so this being optional helped to cater to differing perspectives in the group.

### Facilitators of participation

Flexible and adaptable delivery of the programme, having individual control over movement and sensory input and key logistical considerations were described by participants as central ingredients that promoted programme engagement.

#### Adaptable and flexible delivery

Adaptability was the most important facilitator for participants including lying and sitting options; music being optional; and having sessions at different times of day. Seated or lying down options were particularly useful, allowing participation in the way that best suited participants on any particular day.

> *‘So yeah, like sometimes I go full on full on music, pretending I’m a dancer on stage, and other times I’ve just been lying down with the music. Like at a very minimal volume and just using it more as a meditative and session.’ (Participant 5)*
>
> *‘…the fact that you could just set your own pace as well was amazing. And the ladies always reminded, as you know, if you can’t do the class take it at your own pace, you don’t have to keep up with anyone.’ (Participant 4)*

#### Individual titration of movement and sensory input

Making it clear that the movements could be done however the individual wanted was also very useful, both in ensuring participants did not overexert, but also giving them increased confidence that their own adaptation was correct.

> *‘you don’t need to watch too closely with the instructors are doing. This is a suggestion and you explore the movements that feel comfortable in your own body. So, I think that framing and that kind of repeated invite to to take it slowly. I I think that has helped.’ (Participant 3)*

The ability to reduce sensory inputs to avoid over-stimulation was also valued

> *‘You can even like not listen to the music, and if you’re a bit overstimulated, which is something that I used to have like, especially music or a lot of talking, which kind of quickly got too much….So I really feel like it it is well designed that way that you can kind of adjust the session to what you need to get out of it at the moment.’ (Participant 5)*

#### Simple Logistics

Very simple logistical arrangements and content related to increasing participation was also facilitating for many.

> ‘I think it’s just it’s so easy because they send a link every week…. So, you didn’t have to go back and try and find it, because what I find with technology, I find it difficult to to use technology.’ And ‘you don’t have to remember a yoga mat or a blanket, or, you know, because often you forget all these things and then that makes you stressed and you can’t really enjoy the class.’ (Participant 4)
>
> *‘the repetitiveness of the classes has helped the the cognitive function in in the brain as well, and the breeze in as well.’ (Participant 1)*

Being online enabled people to attend who otherwise would not have been able

> *‘I couldn’t go out to to do a class, so if I didn’t have the online class, I wouldn’t have been participating in anything at all.’ (Participant 4)*

### Barriers to participation

Certain barriers and suggestions for improvement were mentioned by some participants when directly questioned, though all made it clear that these were minor points and should be interpreted in the context of a highly enjoyable and impactful experience. The potential barriers raised were also seen differently by different people, with some seeing things as positive which others saw negatively, while others could see both the positives and negatives regarding a particular component of the programme.

#### Advertising and recruitment

Perhaps the most common negative point raised related to difficulty finding out about the programme. One participant reported searching Google for ballet for long COVID and not being able to find anything. She had previously taken adult ballet classes and was motivated to try the programme after having seen the ENO Breathe programme

> *‘For some reason the Scottish Ballet one didn’t pop up on my searches.’ (Participant 5)*
>
> *‘I really wish I’d known about it a bit earlier because I would have joined the whole programme. But I was surprised there wasn’t more people on it. I think there needs to be more awareness’ (Participant 1)*

#### Online format limited social connectedness

Participants reported that being online made it more difficult to form social connections with other participants,

> *‘So, you know that that’s the only downside I think is the fact that it is it’s on a screen rather than in human but. But I take that any day, even though it is that’s the that’s the negative I would accept that there’s more positive than there is negative from it.’ (Participant 5)*

The majority of participants felt there was little or no impact on their social lives.

> *‘I wouldn’t say that that’s socially, it’s changed really much for me.’ (Participant 4).*

This was largely attributed to the online format, which was not felt to be conducive to forming friendships as such. With the duration of sessions and programme also limiting social bonding of the group

> *‘I think it was probably not the not regular or not long enough for for this to kind of establish something.’ (Participant 3)*

#### Preconceptions of ballet and physical activity

One perceived barrier related to the programme being run by Scottish Ballet, was the perception that the programme was traditional Ballet. Participants commented that this might have put some people off who felt that ‘ballet was not for them’.

Physical activity can be a particularly difficult topic for some people with long COVID. Participants described how discussing the programme with other people with long COVID was not always received positively due to negative experiences of physical activity for some people and the wider discussions around its role in long COVID. This was perceived as a barrier for others to sign up for the programme.

> *‘I’ve tried to encourage a lot of my members to join, but as soon as you say it to them, it’s exercise they’re thinking “I can’t do exercise” and I said, well, I’m living proof of the the movement cause sometimes when I went into my other group I could hardly walk, you know, could hardly, but now I sit up straight. You know, they’re trying to take a better posture and I show them, you know, the exercises that we’re doing and kind of encourage them to try and get involved in these kind of things because I know how good it makes you feel. You know they’re not open to exercise at all, because all they’re reading in the, you know, the World Health Organization “you don’t increase your exercise, you don’t do this because it it’ll always cause your crashes”…. but what they’re not thinking about is it’s not hardly exercise, it’s gentle movement that can be done lying on a bed and it can be done sitting as well and you don’t have to actually stand up and do a lot of movement and things like that. So, I don’t think people realise until they actually start doing something like that, it can be useful for your body.’* (Participant 4)

### Quantitative results

Overall, participants reported improvements to their physical, psychological and social health (figure 1). Improvements to physical and psychological health were attributed to participation (figure 2). Participants felt that their social health was less directly impacted by participation. Six participants felt participation had improved their symptoms to some degree, however seven participants felt the programme had not impacted their symptoms (figure 2). Regarding meeting expectations, eleven said it ‘completely’ and two ‘partially’ met their expectations. Eleven said they would ‘definitely’ continue to use the programme content, one ‘probably’ and one ‘probably not’. All 13 said they would ‘definitely’ recommend the programme to other people with long COVID.

## Discussion

This study explored participant experience of Scottish Ballet’s dance-based long COVID support pilot programme regarding perceived health and wellbeing impacts, and facilitators and barriers to impact. Participants described a range of positive impacts around experience of movement, and consistently reported that they felt the programme had been highly beneficial. Less than half felt the programme had improved their actual symptoms of long COVID, but all reported some degree of positive impact on mental and/or physical health. These impacts came from the direct calming and refreshing experience of the sessions, and a sense of increased resilience related to increasing confidence in movement, creativity, and reengaging with aspects of identity impaired by long COVID. This is an important contribution to work exploring how holistic arts-based interventions can help support people with long COVID.

This study aligns with and builds on other research in this area. Dance-based interventions in other medical conditions have demonstrated potential to deliver holistic impacts often altering the experience of a condition, or a person’s ability to manage the symptoms, rather than the pathophysiological drivers of the disease itself [30, 31, 33–35, 42–44]. The findings also complement other research exploring the use of arts-in-health approaches in long COVID [40, 45] which suggest potential to improve quality of life and symptom management. Being online facilitated the participation of some participants but might have resulted in lower social connection than if it had been in person. Such findings align with other related work using online delivery of an online singing intervention for people with post-natal depression [46], a theatre-based intervention for long COVID [40], and a singing-based online intervention for long COVID[45]. These findings sit within a larger body of research regarding the relative pros and cons of online compared with face-to-face intervention delivery. Commonly identified pros include facilitating access to those not able to attend in person, less potential exposure to infectious disease, reduced time and financial costs[26, 47, 48]. Cons of online delivery include digital exclusion of those with lower levels of digital literacy or lacking required equipment, concerns regarding security and privacy, and reduced sense of social connection[26, 45, 47, 48].

The themes identified include both programme ingredients and mechanisms which interact with each other. For example, participants valued spending time in a ‘safe space’ as an end in itself and a pleasant experience, but this also triggered a mechanism of increased confidence in movement. As with other complex interventions, various, dynamic, and interacting mechanisms appear to be occurring[49]. The INNATE checklist in the supplement provides more detailed information aspects of the project, people, and context that may be contributing to perceived health impacts. Key mechanisms that were important here include low intensity dance-based movement in a calm and supported environment, where the participant felt in control and was able to build confidence in movement. This important relationship of movement within music or dance has also been described in relation to other interventions [50, 51]. Mindful movement creating feelings of calm and connection with self was another key mechanism. Systematic reviews and metanalyses of interventions using mindful movement suggest that such approaches lead to improvements in mental health and quality of life, however the quality of studies included is frequently poor, hence firm conclusions are difficult to make [52, 53]. Having shared lived experience was an important source of social connection, with both contributing to the calm and calming environment experienced. These elements have also been identified in other research as being central to supporting coping for people with long COVID [11, 54].

### Considerations and limitations

Certain considerations and limitations to this study should be discussed. Firstly, this is a small study, and therefore the generalisability remains unclear. Secondly, it is likely that those interviewed represent a self-selected group of individuals who feel that they benefited from participation. Those that felt it was not useful are likely to have stopped attending or might not have volunteered to participate in the evaluation. Similarly, it is likely that certain people with long COVID did not participate due to specific aspects of their condition making the programme unsuitable for them. For example, people with very severe post exertional symptom exacerbation.

The programme clearly had a positive impact on the individuals who participated in the evaluation; however, other people who might have liked to participate might not have been aware of the sessions, which was highlighted as a barrier by participants. The challenge is how best to identify and contact such individuals. Future work should include wider dissemination to reach more potentially interested people living with long COVID. Discussion with the pilot participants regarding specific long COVID support groups, networks, and existing service providers would be a logical place to start.

Given the size of the sample it was not possible to employ additional more rigorous methods to assess impact, such as conducting a clinical trial, with randomisation, blinding, and quantitative assessment of change with validated measures. However, the current evaluation provides a strong foundation for future work.

## Conclusions

The Scottish Ballet’s long COVID support programme pilot was well received by participants who reported positive holistic impacts, related to building confidence in movement and developing ways to manage symptoms. This pilot work has demonstrated potential to further develop and expand the programme. Long COVID remains a major issue for those with the condition, and society more broadly, with limited management options. There is clearly a need for holistic approaches to support people with long COVID, alongside ongoing efforts to improve understanding of the condition and develop effective, evidence-based interventions.

The programme appears to provide something unique and highly valued by participants. While the programme might not be suitable to everyone with long COVID, for self-selecting individuals, this programme has the potential to make a meaningful contribution. The findings of this evaluation will be useful to inform Scottish Ballet’s understanding of the impact of their Health programmes on participants, as well as shaping the programmes development and future application.

## Supporting information

Reporting checklist

## Data Availability

Data from this study are not being made available for sharing, as given the relatively small number of participants, it would not be possible to anonymise interview transcripts sufficiently.

## Author Contributions

KEJP led the study including design with AB and NSH. KEJP and NSH gained the ethical approval. KEJP conducted and transcribed the interviews. KEJP, AB, AL and SB conducted the analysis. KEJP wrote the first draft of the manuscript. All authors (KEJP, AB, AL, SB, PW, MIP, DF, NSH) contributed to the study design, writing, reviewing and editing the manuscript, and approved the final manuscript for submission.

## Reporting Checklist

See attached COREQ checklist.

## Acknowledgements

We the authors would like to thank the study participants. KEJP was supported by the Imperial College National Heart and Lung Institute Clinical Lecturer. KEJP would like to acknowledge the National Institute for Health Research (NIHR) Biomedical Research Centre based at Imperial College Healthcare NHS Trust and Imperial College London for their support. The views expressed are those of the authors and not necessarily those of the NHS, the NIHR, or the Department of Health.

## Data availability statement

Data will not be made publicly available as anonymisation would not be possible.

## Contributors

KEJP, AB and NSH designed the study. KEJP conducted the interviews, led the analysis and wrote the first draft. AB, AL, SCB, PJW also contributed to analysing the data. All authors contributed to interpretation of the results, commenting on drafts and revisions of the manuscript, and agreed on the final draft for submission.

